# Muscle Synergies for Post-Stroke Motor Assessment and Prediction in a Randomized Acupuncture Trial

**DOI:** 10.1101/2024.01.28.24301900

**Authors:** Fung Ting Kwok, Ruihuan Pan, Shanshan Ling, Cong Dong, Jodie J. Xie, Hongxia Chen, Vincent C. K. Cheung

## Abstract

Motor rehabilitation after stroke is challenging due to the heterogeneity of stroke presentations. Maximizing recovery hinges on suitable personalization of rehabilitation, which depends on reliable motor assessments and predictions of the subjects’ responses to different interventions using biomarkers of brain impairment. Beyond the previously defined neuroimaging biomarkers, impairment-dependent motor patterns of stroke survivors during voluntary movement are alternatives that potentially offer accurate and precise predictions. Specifically, muscle synergies identified from multi-muscle electromyographic signals (EMG), as neuromotor modules employed by CNS for muscle coordination, have been previously used to evaluate upper limb functions post-stroke in small-to-moderate-sized cohorts. While these initial results appear promising, how muscle synergies should be most profitably used for clinical assessments and whether they predict post-rehabilitation responses remain unexplored.

Here, we evaluate the potential of muscle synergies in assessing upper limb motor functions and predicting outcomes from multiple rehabilitative options in a moderately large cohort of subacute stroke survivors (N=88, 55±35 days post-stroke) recruited for a randomized trial of acupuncture as an adjunctive rehabilitative intervention. Subjects (N=59) were randomly assigned to monthlong abdominal acupuncture (Acu), sham acupuncture (ShamAcu), or no acupuncture (NoAcu), alongside basic care. Four clinical scores and EMGs of the stroke-affected upper limb (14 muscles, 8 tasks) were collected before and after intervention. Muscle synergies were extracted from EMGs using factorization. For each subject, features of the synergies and their temporal activations were comprehensively summarized by 12 muscle synergy indexes (MSI).

We first demonstrated cross-sectionally that our MSIs correlated significantly with all clinical scores, and thus could capture impairment-related synergy changes. Longitudinally, Acu was differentiated from ShamAcu and NoAcu in having clinical score improvements accompanied by the restorations of more MSIs. For each treatment group, we then built regression models that predict clinical scores’ realized recovery from pre-intervention MSIs and other variables. Model-predicted recovery correlated significantly with observed recovery (R^2^=0.53-0.70). To test the models’ utility in patient stratification, for every Acu and NoAcu subject we retrospectively identified the intervention option expected to yield greater recovery by comparing the predicted Acu and NoAcu outcomes. Subjects who indeed received the model-assigned intervention showed more realized recovery in Fugl-Meyer Assessment (section A) than those who received incorrectly assigned intervention (p=0.013). Overall, our findings suggest that muscle synergies, when suitably summarized as MSIs, may clarify the intervention’s effects and assist in motor assessment, outcome prediction, and treatment selection. MSIs can be useful recovery biomarkers in future schemes of precision rehabilitation.

## INTRODUCTION

Stroke is a prevalent neurological disorder and a leading cause of adult disability. It occurs when blood flow to the brain is disrupted by a blockage (ischemic stroke) or rupture (hemorrhagic stroke).^1^ Since motor dysfunction is a common complication of stroke, motor rehabilitation is crucial for restoring quality of life post-stroke. Unfortunately, the development of new, effective stroke rehabilitation has been challenging.^2^ Given the heterogeneity of stroke presentations,^3^ maximizing post-rehabilitation motor recovery hinges on suitable personalization of intervention, which in turn depends on reliable assessments of the motor system and accurate predictions of each survivor’s responses to different interventions. Such assessments and predictions for recovery may be aided by biomarkers.^4^ Defining clinically effective recovery biomarkers remains an important question in stroke rehabilitation.^5^

The ideal recovery biomarker should presumably align well with the neurological principles of motor control and the structure of the motor system.^6,7^ It has been demonstrated that the motor system is organized into discrete modules, called muscle synergies, whose flexible combination generates diverse movement patterns^8,9^ (Supplementary Note 1). The brain likely coordinates movements by activating the groups of muscles organized within the muscle synergies, as opposed to individual muscles, to reduce the computational burden of control. Muscle synergies may be conveniently identified from behavioral multi-muscle surface electromyographic data (EMG) through factorization.^10^ In contrast to information from electroencephalography and neuroimaging modalities, muscle synergies reflect the low-level control architecture of the motor system down at the level of multi-muscle coordination. Thus, muscle synergies have been proposed as recovery markers that may also be targets of rehabilitative interventions^11–13^ and potentially useful for post-stroke motor assessments^11,12,14–16^ and predictions of recovery.^17^ Previous studies on relating synergies to motor outcomes have developed numerous muscle synergy indexes (MSIs) to quantify the characteristics of muscle synergies and their corresponding activation profiles.^18–21^ These MSIs, categorizable into 6 types depending on the synergy features they summarize (Supplementary Table 1), were then correlated with motor functional and clinical scales across stroke survivors.

The MSIs can be useful for stroke rehabilitation in the following ways. First, MSIs can supplement clinical scores such as the Fugl-Meyer Assessment in evaluating post-stroke motor deficits. Clinical scores are advantageous because they can be assessed quickly with reasonable reliability, but they convey limited information on how any therapy may modify the motor system as functional gain is achieved. Some clinical scores only reflect the degree of task completion but disregard how exactly the tasks are carried out,^22^ thus they cannot determine whether post-training improvement is due to the true recovery of normal motor patterns or the emergence of new compensatory patterns. The MSIs may differentiate the two scenarios, in that for any functional improvements suggested by clinical scores, true recovery is indicated if MSIs change towards their normative levels, and compensation if otherwise. Second, as indexes based on motor modules with a concrete neural basis, MSIs are potentially useful for predicting the outcome of rehabilitation. This is especially so given the predictive successes of previous neurophysiological and neuroimaging markers (Supplementary Table 2), such as those employed in the predict recovery potential (PREP) algorithm.^23^

In this study, we seek to systematically evaluate a comprehensive set of MSIs in terms of their assessment and predictive potential in stroke rehabilitation. While previous MSI studies typically included 2 to 5 MSIs from up to 4 MSI categories, here we revised and expanded the prior repertoire of MSIs to include 12 MSIs summarizing diverse muscle synergy features (all 6 categories in Supplementary Table 1). Also, while previous MSI investigations on neurological diseases recruited an average of 15 patients,^16^ we recruited a moderately large sample (n = 88) of stroke survivors with pre- and post-treatment recordings from 59 of them. Our inclusion of more MSIs and a greater number of subjects has enabled us to not only evaluate the MSIs’ potential for motor assessment, but also formulate MSI-based treatment-specific predictive models to test, retrospectively, whether assigning each subject to a treatment option based on model predictions would lead to an overall better rehabilitative outcome.

Our MSI evaluation was performed in a randomized clinical trial designed to evaluate the effectiveness of abdominal acupuncture^24,25^ in promoting motor recovery in subacute stroke survivors. The choice of acupuncture for our evaluation is suitable for the following reasons. First, the acceptance of acupuncture as an adjunctive therapy for stroke rehabilitation has increased worldwide in recent decades.^26,27^ Second, despite evidence from both human^25,28^ and animal^29^ studies demonstrating that acupuncture is beneficial for survivors of ischemic stroke, it is unknown whether it promotes true recovery or compensation. Our MSI analysis may provide insights on this question. Third, although whether the outcome of acupuncture was good or poor could be predicted by the extent of lesion in the periventricular white matter,^30^ this prediction is dichotomous and therefore has limited use in precise patient stratification, especially if one intends to compare the prediction of acupuncture with those of other therapies in order to determine the optimal option for a stroke survivor. Thus, we also ask whether MSIs could predict a continuous outcome variable for stroke rehabilitation.

## MATERIALS and METHODS

### Subject recruitment

A total of 88 first-onset stroke survivors (age = 56.23 ± 10.95 years [mean ± SD]; female = 31; post-stroke duration = 55.01 ± 34.51 days) with upper-limb hemiparesis (Supplementary Table 3) were recruited from the Guangdong Provincial Hospital of Chinese Medicine. Among them, 59 participated in a prospective, sham-controlled, randomized clinical trial (trial no. NCT03712085 at ClinicalTrials.gov) and received acupuncture (Acu) (n = 21), sham acupuncture (ShamAcu) (n = 21), or no acupuncture (NoAcu) (n = 17). Pre-intervention demographic and clinical data are summarized in Supplementary Table 3. A CONSORT flow diagram for subject recruitment and study design is included in Supplementary Figure 1. Also, 10 adults with no neurological disorders (age = 28.00 ± 4.92; female = 6) were recruited as control subjects. All stroke survivors and control subjects were right-handed. All participants gave informed consent prior to experimentation.

Initial inclusion criteria of the stroke survivors were: (1) First-onset stroke survivors with hemiplegia, with diagnosis of left or right cerebral infarction in the middle cerebral artery supply area confirmed by brain CT or MRI; (2) Aged between 35 and 75 years old; (3) Stroke survivors at 0.5 to 3 months post-stroke presented with stable vital signs; (4) No severe aphasia and cognitive impairment, and were able to understand and execute commands; (5) Able to control sitting balance without external support, with the Brunnstrom stage of hemiplegia (upper limb and hand) being II or above; (6) Agreed to sign the informed consent; (7) The ethics committee agreed to the subject’s participation. To enable a greater number of patients to partake in the study, the initial inclusion criteria were modified one year after the study began. Item 1 was amended to include bilateral cerebral infarction, and item 5 was changed to Brunnstrom stage II or higher. Exclusion criteria included: (1) History of recurrent stroke, subarachnoid hemorrhage, or brain tumor; (2) Contraindication to undergo 3T MR imaging; (3) Claustrophobia; (4) History of severe complications involving the heart or any hepatic or renal diseases; (5) History of non-compliance with medical interventions; (6) Had participated in other clinical trials recently. Both the original and revised research protocols were approved by the Ethics Committee of Guangdong Provincial Hospital of Chinese Medicine (initial: NO. BF2018-164-01, revised: NO. BF-2018-164-02).

The sample size of this study was calculated based on a previous clinical report^31^ with the MATLAB function sampsizepwr. Assuming the FMA score is 30 ± 10 prior to treatment and 37 subsequent to treatment, the effect size between groups is 0.7. For a statistical power of 0.8, a minimum of 19 cases per group was required. Considering the uncontrollable circumstances that could result in a 15% subject dropout rate, 3 additional subjects were added to each group. Thus, at least 22×3 = 66 subjects needed to be enrolled in the designed randomized controlled trial (RCT). After excluding the lost follow-up subjects, 59 subjects were finally enrolled in the RCT trial. Additionally, to enlarge the baseline EMG dataset of stroke patients, we recruited 32 patients who met the inclusion criteria after the closure of the RCT and completed the EMG and basic clinical assessment before intervention.

### Blinding and interventions

Eligible patients were randomly divided into 3 groups in a 1:1:1 ratio following a random number table. For the concealment of allocation, the grouping was decided by the subject number used in the random-number list. The matching between the subject number and group assignment was then concealed, and only revealed to the acupuncturist right before the delivery of the intervention. The physicians responsible for evaluating the clinical scores and collecting surface electromyographic (EMG) activities were blinded to the grouping.

All interventions lasted for one month. The NoAcu subjects were notified that they would not receive acupuncture when they signed the consent form, while the Acu and ShamAcu subjects were not informed of their treatment assignment. The Acu and ShamAcu groups received the therapeutic and sham versions of Bo’s abdominal acupuncture, respectively, with a dosage of 30 minutes per day, 5 days per week,^24,32^ alongside basic care. The NoAcu group received basic care only. Basic care consisted of conventional upper-limb rehabilitative training and measures for secondary stroke prevention. For the Acu group, abdominal needles (0.2 x 30 mm; Suzhou Hualun Medical Appliance Co., Ltd., China) were inserted into eight abdominal acupuncture points, including Guanyuan (RN4), Qihai (RN6), Xiawan (RN10), Zhongwan (RN12), ipsilateral Shangqu (KI17), ipsilateral Huaroumen (ST24), ipsilateral Shangfengshidian (AB1), and ipsilateral Shangfengshiwaidian (AB2). For the ShamAcu group, customized flat needles (0.3 x 30 mm; Changzhou Dayi Medical Device Co., Ltd., China) were inserted into the same acupoints to produce a placebo effect.

### Clinical assessments

For all stroke survivors in the RCT trial (n = 59), clinical assessments were conducted at weeks 0 (pre-intervention), 2 (half-completed intervention), and 4 (post-intervention). We selected multiple clinical assessments from both the body function/impairment and activity/disability domains under the International Classification of Functioning, Disability and Health (ICF),^33^ including the Fugl-Meyer Assessment of the Upper Extremity (FMA(UE)) and the Brunnstrom Stage (BS) for the former domain, and the Wolf Motor Function Test (WMFT) and the modified Barthel Index (BI) for the latter. Since we also recorded surface EMG activities of shoulder-arm muscles (see below), we found it useful to calculate two additional clinical scores, FMA(A) and BI(UE), by selecting a subset of task items from FMA(UE) (section A) and BI (items 1-6), respectively. These selected items specifically reflect the functions of the recorded muscles. The task items used for all clinical assessments are listed in Supplementary Table 4.

For all clinical scores of each RCT subject, we calculated both their absolute changes after intervention and their realized recovery, defined as the actual score change relative to the maximum possible score improvement.^34,35^ Thus,

> Realized Recovery = (Score_post_ – Score_pre_) / (Maximum Score – Score_pre_).

For the rest of the stroke survivors not in RCT (n = 29), FMA(UE) and BI were evaluated upon recruitment.

### EMG recording and muscle synergy extraction

We recorded EMGs from 14 shoulder, arm, and back muscles of the stroke-affected upper limbs (Supplementary Table 5) of stroke survivors at weeks 0 and 4, along with EMGs from each of both upper limbs of healthy subjects, which served as a baseline for comparison. Specifically, we recorded 167 EMG sessions, including 88 pre-intervention sessions (for all stroke survivors) and 59 post-intervention sessions (for stroke survivors in the clinical trial), as well as 20 sessions from healthy subjects (10 sessions for each side). During EMG recording, the subject performed 8 upper-limb motor tasks related to activities of daily living, including those described in Cheung et al.^14^ and other tasks (Supplementary Table 5), with each task repeated 5 times. All EMGs were preprocessed with filtering, rectification, integration, and variance normalization as described in Cheung et al.^36^

For every EMG session, we used nonnegative matrix factorization (NNMF)^37^ to decompose the EMG (**D**) into time-invariant muscle synergies (**W**) and their activation profiles (**C**) (Supplementary Note 2). To reflect the global data structures, we first extracted the overall muscle synergies from the EMGs of all tasks, with the dimensionality identified as the minimum number of muscle synergies that attained an R^2^ > 0.80 in *both* the all-task EMG and each of the 8 single-task EMGs. To reflect task-specific data structures, we then extracted task-specific muscle synergies from the EMG of each task, with the dimensionality identified as the minimum number of synergies that yielded an R^2^ > 0.80 for the single-task EMG. The R^2^ value was defined as in Cheung et al.^36^ For each NNMF decomposition, the synergy extraction procedure was repeated 50 times; each repetition was terminated when the change in R^2^ was less than 0.001% for 20 consecutive iterations.

To characterize preservation, merging, and fractionation of muscle synergies after stroke,^14^ we categorized the muscle synergies of stroke survivors and healthy subjects into post-stroke and healthy synergy clusters, respectively, using k-means (Supplementary Fig. 2). For each number of clusters, k-means clustering was conducted 10,000 times. The optimal number of clusters was defined as the minimum number of clusters at which each subject had no more than one muscle synergy represented in the same cluster.

### Computation of muscle synergy indexes

We designed 12 muscle synergy indexes (MSIs) (names and abbreviations listed in Table 1) to quantify the characteristics of the muscle synergies and their activation profiles. The rationale and detailed computation procedure for each MSI are described in Supplementary Note 3. Briefly, indexes DevD_O_ and DevD_A_ represent the deviation of the post-stroke numbers of muscle synergies from the normative; BFRR_W_, BFRR_C_, and BFRR_C_ (mod) assess the normalcy of the **W** and **C** matrices after stroke; MI and FI quantify post-stroke muscle synergy merging and fractionation, respectively^14^ (Supplementary Table 6); DO and MEA calculate the extent of oscillation and the magnitude of activation of the activation profile, respectively; and ITV_BFRR_W_, ITV_BFRR_C_, and ITV_BFRR_C_ (mod) determine the variability of the **W** and **C** matrices across tasks.

**Table 1.** Names and abbreviations of the 12 muscle synergy indexes (MSIs). The rationale and detailed computational procedure for each MSI are included in Supplementary Note 3.

| Abbreviation | Full name |
| --- | --- |
| 1) Dimensionality of the the impaired limb |  |
| DevD<sub>O</sub> | Deviation in the Dimensionality from normal (original value) |
| DevD<sub>A</sub> | Deviation in the Dimensionality from normal (absolute value) |
| 2) Similarity of the W matrix between the impaired limb and the reference limb |  |
| BFRR<sub>W</sub> | Bidirectional Fitting R<sup>2</sup> Ratio (BFRR) of the W matrix |
| 3) Other features of the W matrix |  |
| MI | Merging Index |
| FI | Fractionation Index |
| 4) Similarity of the C matrix between the impaired limb and the reference limb |  |
| BFRR<sub>C</sub> | Bidirectional Fitting R<sup>2</sup> Ratio (BFRR) of the C matrix |
| BFRR<sub>C</sub> (mod) | Bidirectional Fitting R<sup>2</sup> Ratio (BFRR) of the C matrix (modified) |
| 5) Other features of the C matrix |  |
| DO | Degree of Oscillation of the C matrix |
| MEA | Magnitude of Effective Activation of the C matrix |
| 6) Inter-task variability in the W or C matrix of the impaired limb |  |
| ITV_BFRR<sub>W</sub> | Inter-Task Variability (measured by BFRR) in the W matrix |
| ITV_BFRR<sub>C</sub> | Inter-Task Variability (measured by BFRR) in the C matrix |
| ITV_BFRR<sub>C</sub> (mod) | Inter-Task Variability (measured by BFRR) in the C matrix (modified) |

While some of the MSIs were computed solely by using the **W**s and **C**s of the stroke-affected limbs (DO, MEA, ITV_BFRR_W_, ITV_BFRR_C_, ITV_BFRR_C_ (mod)), some others (DevD_O_, DevD_A_, BFRR_W_, BFRR_C_, and BFRR_C_ (mod)) were computed by comparing the **Ws** and **Cs** of stroke survivors with those of healthy subjects. Specifically, for the latter group of MSIs, the affected limb of each stroke survivor was compared with the side-matched healthy limbs (e.g., if the left side was affected, the affected limb was matched with the left limbs of healthy subjects). For these MSIs, the same computation was performed by taking each side-matched healthy limb as baseline, and the final MSI value was calculated as the mean across the values derived from all healthy limbs. MI and FI were computed by comparing the **W** of each stroke survivor with the centroids of healthy synergy clusters.

### Statistics analyses

All data analysis was performed in Matlab (R2022a). We used the Lilliefors test to evaluate if a sample followed a normal distribution. For paired samples, we assessed group differences by the paired t-test (for normal samples) or Wilcoxon signed-rank test (for non-normal samples). For independent samples, we assessed group differences by the independent t-test or one-way ANOVA (for normal samples), or the Mann-Whitney U test or Kruskal-Wallis (KW) test (for non-normal samples). If the result of the ANOVA or KW test was significant, we performed a post-hoc Tukey-Kramer multiple comparison to determine the significance of each group combination.

Pearson correlation was used to assess the linear relationship between two variables. Multiple linear regression was used to determine how much of the variance of the dependent variable could be accounted for by multiple independent variables. Stepwise multiple linear regression (p_enter_ = 0.05, p_remove_ = 0.10) was used to select the appropriate input variables that contribute to the prediction of the output variable.

## RESULTS

This study adopted a combined cross-sectional-longitudinal design. We recruited 88 stroke survivors, 59 of whom, following the original allocation, participated in a one-month clinical trial of acupuncture between January 2019 and May 2022. We first used the pre-intervention data of all stroke survivors (n = 88) to explore how muscle synergies changed after stroke and the relationship between the MSIs and clinical scores. Then, we used the pre- and post-intervention data of those in the clinical trial (n = 59) to investigate the effect of acupuncture on post-stroke motor recovery, and evaluate the effectiveness of the MSIs in predicting rehabilitation outcomes.

### MSIs captured impairment-relevant muscle synergy changes post-stroke

We began our investigation by visually comparing the muscle synergies and activation profiles of severely impaired stroke survivors (FMA(A) ≤ 10) with those of healthy subjects (Fig. 1). For example, when examining the muscle synergies of subjects S11 (FMA(A) = 6/36) and H12 (healthy), we observed that the severely impaired had a greater number of muscle synergies, which were also more fragmented (i.e., sparser in representation) (Fig. 1B). In addition, the activation profiles of the severely impaired appeared more oscillatory and irregularly shaped in time, and had a smaller area under the curve, especially when the oscillation area was disregarded (Fig. 1C). In another pair of stroke survivor (S25, FMA(A) = 9/36) and healthy subject (H1), we examined which synergies were activated in which tasks for both subjects (Fig. 1D). It is apparent that there were more varieties of synergy selections deployed across tasks in the healthy subject than in the stroke survivor. The above synergy differences are described in more detail in Supplementary Note 4.

**Figure 1.**
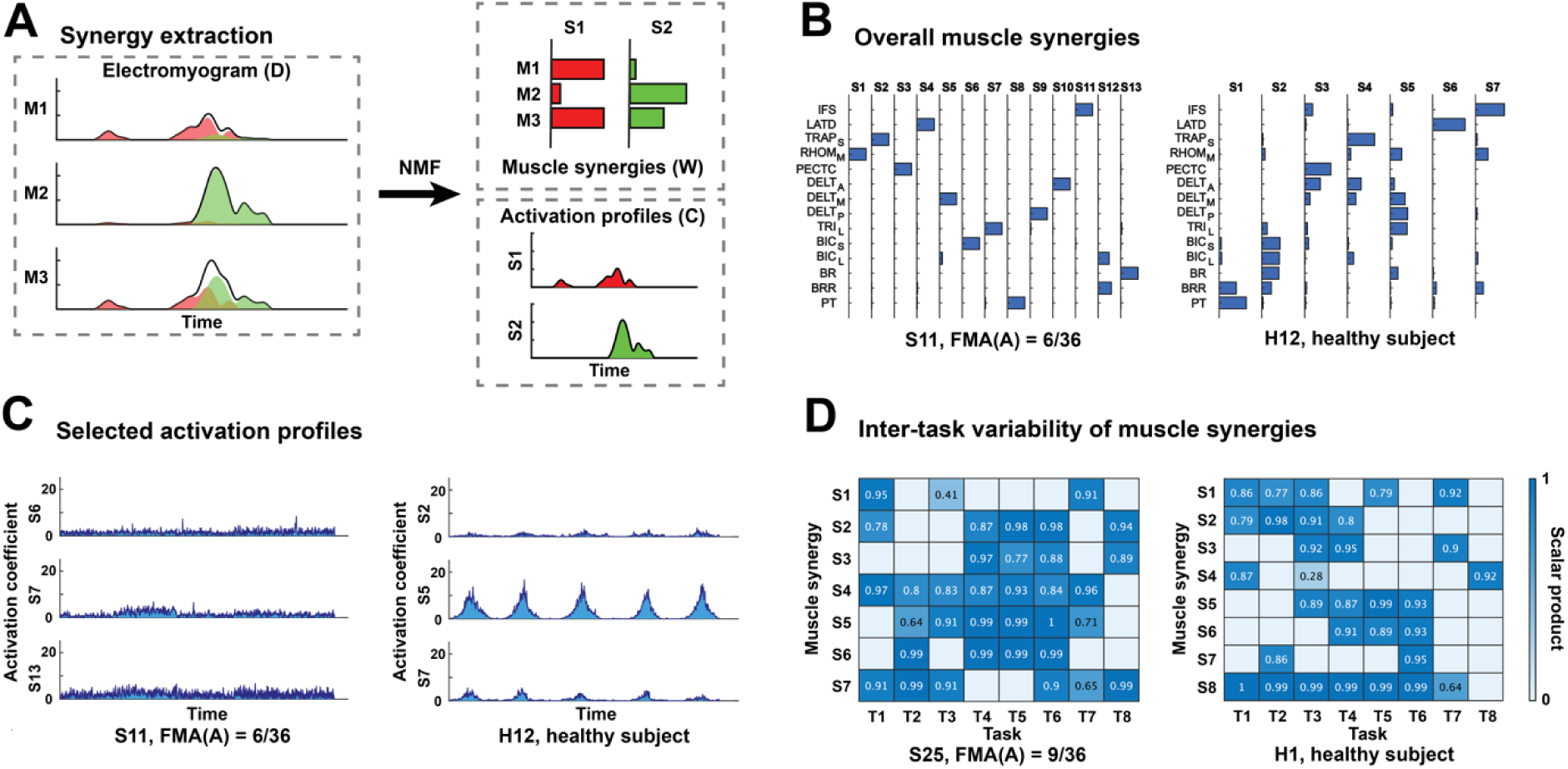
Post-stroke alterations in muscle synergies and activation profiles. (A) A schematic of synergy extraction. The preprocessed EMG of every session was decomposed into muscle synergies (i.e., the W matrix) and activation profiles (i.e., the C matrix) using non-negative matrix factorization (NMF). (B) The overall muscle synergies of a severely impaired stroke survivor (S11, FMA(A) = 6/36) and a healthy subject (H12). S11 had a greater number of muscle synergies, and the synergy compositions were more fragmented. (C) The selected activation profiles (i.e., those of the three most activated synergies in task 8) of the same pair of subjects. The activation profile of S11 exhibited more oscillations, more irregularity, and a smaller area under curve. (D) The inter-task variability of muscle synergies of another pair of severely impaired stroke survivor (S25, FMA(A) = 9/36) and healthy subject (H1). Each task-specific muscle synergy was matched with an overall muscle synergy of the same subject on a one-to-one basis. The numbers on the colormap represent the scalar products between each task-specific synergy and the matched overall synergy. Severely impaired stroke survivors tended to employ the same set of synergies across tasks. For S25, synergies 1, 4, 5, and 7 were employed in both task 3 and task 7, while synergies 2 – 6 were employed in tasks 4 – 6.

Motivated by the above observations, we developed 12 MSIs to quantify the muscle synergy differences between the stroke survivors and healthy subjects (Table 1; Supplementary Note 3) and computed the MSIs for all subjects in both groups. For all MSIs except the merging index (MI), values from the severely impaired stroke survivors were significantly different from their respective baselines from healthy subjects (p < 0.05). The Pearson correlation between the MSIs and the clinical scores also showed significant correlations (p < 0.05) in 57 out of 60 (from 12 MSIs × 5 scores) instances (Fig. 2A). These results suggest that our MSIs were able to capture the post-stroke muscle synergy changes that are relevant to the severity of functional and motor impairment.

**Figure 2.**
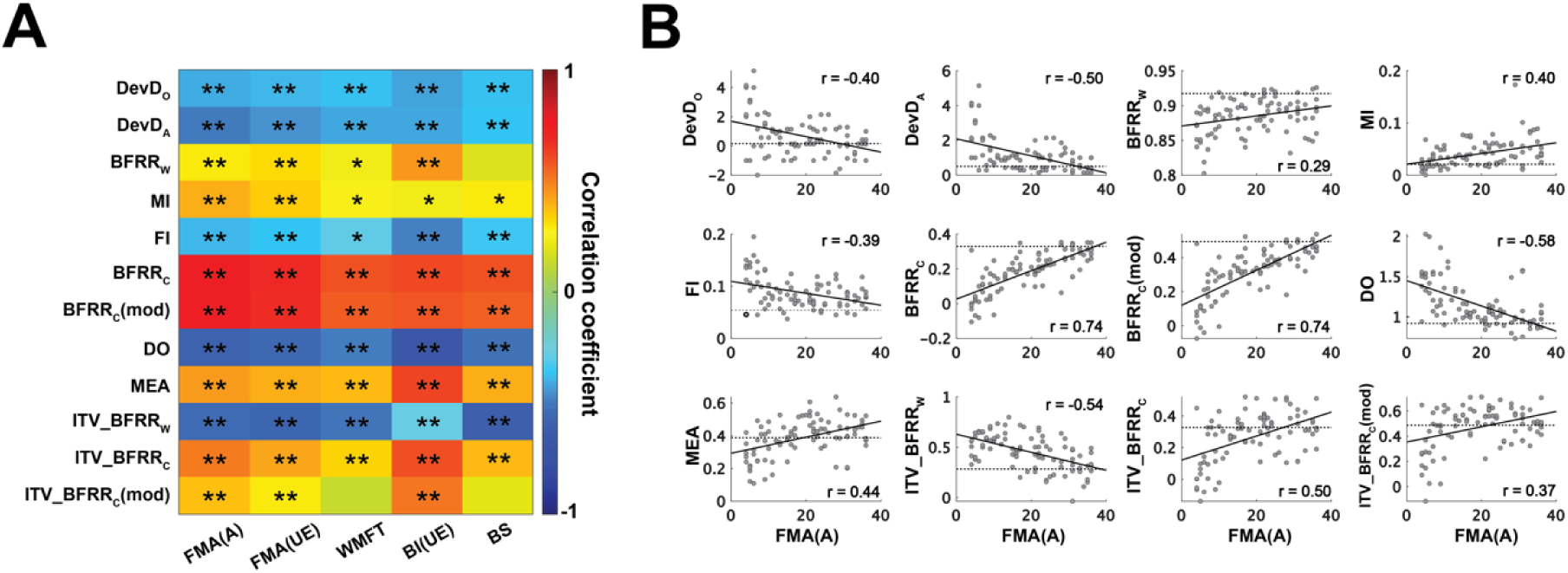
Muscle synergy indexes (MSIs) are impairment-relevant. (A) Pearson correlation between 12 muscle synergy indexes (MSIs) and 5 clinical scores at week 0. *: p < 0.05; ** p < 0.01. (B) Scatterplots of MSIs and FMA(A). The dashed line in each scatterplot represents the median baseline value for each MSI.

As examples, we show in Fig. 2B scatterplots of the 12 MSIs against the FMA(A) of all stroke survivors. We note that MI behaved differently from the other MSIs. For all other MSIs, the difference between the post-stroke and baseline MSIs was the greatest at low clinical scores, as expected. But for MI, the difference was minimal at low clinical scores and maximal at high clinical scores.

### Acupuncture promoted more restorations of MSIs than control interventions

We investigated the effect of acupuncture on post-stroke motor recovery by analyzing its impact on clinical scores and MSIs. We calculated both the absolute change in a clinical score, and the realized recovery of a clinical score by dividing the change in score by the maximum possible score improvement.^34,35^ Across the Acu, ShamAcu, and NoAcu groups, there was no significant group difference in the change in clinical scores or their realized recovery at both weeks 2 and 4 (p > 0.10) except for score BS (Table 2). For this score’s realized recovery, at week 2, the Acu group achieved significantly more recovery (median = 0.14) than the ShamAcu (median = 0) and NoAcu (median = 0) groups (KW test: Chi-sq(2) = 6.83, p = 0.033).

**Table 2.** Rehabilitation outcomes of the Acu group (n = 21), ShamAcu group (n = 21), NoAcu group (n = 17), and all RCT participants (n = 59). For the clinical scores and muscle synergy indices (MSIs), we compared the pre-intervention (week 0) and post-intervention (week 4) values of each group, and for the clinical scores only, pre-intervention (week 0) and half-completed-intervention (week 2) values, by paired t-test or Wilcoxon signed-rank test († p < 0.08; * p < 0.05; ** p < 0.01). The values listed in the table are the average changes across the subjects in each group (mean ± SD). Additionally, we compared the 2- or 4-week changes of the three interventions by one-way ANOVA or Kruskal-Wallis test. The clinical scores and MSIs showing statistically significant or nearly significant inter-group differences are highlighted in bold in the first column (^ p < 0.06; # p < 0.05).

|  | Acupuncture | Sham<br>acupuncture | No acupuncture | All groups |
| --- | --- | --- | --- | --- |
| Change in clinical scores (mean ± SD) after 2 weeks | | | | |
| FMA(A) | 2.05 ± 2.19** | 2.62 ± 3.05** | 1.41 ± 1.50** | 2.07 ± 2.43** |
| FMA(UE) | 3.95 ± 5.14** | 3.62 ± 4.62** | 2.82 ± 2.28** | 3.51 ± 4.33** |
| WMFT | 5.62 ± 5.07** | 4.71 ± 4.69** | 3.35 ± 2.89** | 4.64 ± 4.49** |
| BI(UE) | 5.43 ± 5.20** | 3.29 ± 5.41* | 2.88 ± 3.61** | 3.93 ± 5.00** |
| **BS#** | 0.86 ± 0.83** | 0.57 ± 0.73** | 0.35 ± 0.84 | 0.61 ± 0.82** |
| Change in clinical scores (mean ± SD) after 4 weeks | | | | |
| FMA(A) | 4.62 ± 4.33** | 5.14 ± 4.80** | 5.71 ± 6.18** | 5.12 ± 5.11** |
| FMA(UE) | 9.76 ± 8.27** | 8.76 ± 7.32** | 9.94 ± 6.68** | 9.46 ± 7.52** |
| WMFT | 12.05 ± 8.40** | 10.33 ± 8.71** | 7.47 ± 5.13** | 10.12 ± 7.94** |
| BI(UE) | 10.86 ± 8.29** | 6.24 ± 7.65** | 8.41 ± 6.80** | 8.51 ± 7.90** |
| BS | 1.52 ± 1.22** | 1.24 ± 1.23** | 1.29 ± 1.45** | 1.36 ± 1.30** |
| Change in muscle synergy indexes (mean ± SD) after 4 weeks | | | | |
| DevD<sub>O</sub> | -0.81 ± 1.56* | 0.19 ± 1.30 | 0.00 ± 0.97 | -0.22 ± 1.39 |
| DevD<sub>A</sub> | -0.21 ± 1.30 | -0.15 ± 0.90 | 0.07 ± 0.76 | -0.11 ± 1.03 |
| BFRR<sub>W</sub> | 0.00 ± 0.03 | 0.00 ± 0.02 | -0.01 ± 0.03 | 0.00 ± 0.03 |
| **MI#** | 0.01 ± 0.02* | -0.01 ± 0.03 | 0.00 ± 0.03 | 0.00 ± 0.03 |
| **FI#** | -0.02 ± 0.03* | 0.00 ± 0.03 | 0.01 ± 0.04 | 0.00 ± 0.04 |
| BFRR<sub>C</sub> | 0.04 ± 0.09* | 0.02 ± 0.07 | 0.03 ± 0.06 | 0.03 ± 0.07** |
| BFRR<sub>C</sub> (mod) | 0.05 ± 0.12† | 0.03 ± 0.08 | 0.03 ± 0.08 | 0.04 ± 0.10** |
| DO | -0.12 ± 0.25* | 0.03 ± 0.20 | 0.03 ± 0.20 | -0.02 ± 0.23 |
| **MEA^** | 0.04 ± 0.11 | -0.02 ± 0.08 | -0.03 ± 0.08 | 0.00 ± 0.10 |
| ITV_BFRR<sub>W</sub> | -0.03 ± 0.16 | -0.06 ± 0.13† | 0.00 ± 0.15 | -0.03 ± 0.15 |
| ITV_BFRR<sub>C</sub> | 0.06 ± 0.16 | 0.00 ± 0.12 | 0.05 ± 0.10* | 0.04 ± 0.13* |
| ITV_BFRR<sub>C</sub> (mod) | 0.07 ± 0.19 | -0.01 ± 0.14 | 0.06 ± 0.10* | 0.04 ± 0.16† |

For the MSIs, statistically significant group differences were found for the 4-week changes in MI, FI, and MEA (Fig. 3, Table 2). For MI change, the Acu group (mean = 0.01) had a greater increase than the ShamAcu (mean = -0.01) and NoAcu (mean = 0) groups (one-way ANOVA: F(2,56) = 3.40, p = 0.040). For FI change, Acu (mean = -0.02) had a greater reduction than ShamAcu (mean = 0) and NoAcu (mean = 0.01) (one-way ANOVA: F(2,56) = 3.34, p = 0.043). Similarly, for MEA change, Acu (mean = 0.04) had a greater increase than ShamAcu (mean = -0.02) and NoAcu (mean = -0.03) (one-way ANOVA: F(2,56) = 2.99, p = 0.058).

**Figure 3.**
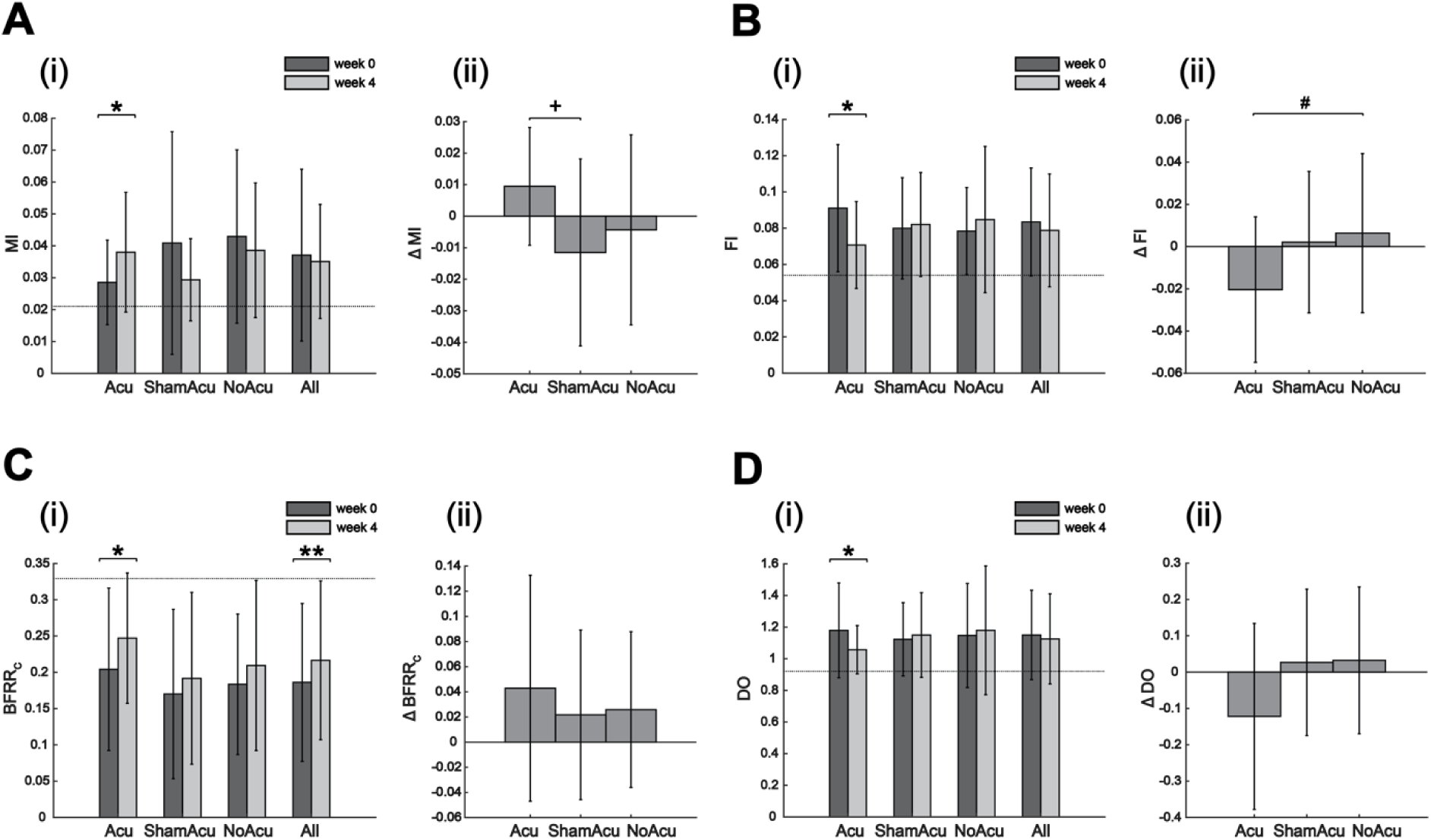
Acu group (n = 21) experienced more changes in muscle synergy indexes (MSIs) than the ShamAcu (n = 21) and NoAcu (n = 17) groups, particularly in (A) MI, (B) FI, (C) BFRR_C_, and (D) DO. For each MSI, we (i) compared the pre-intervention (i.e., week 0) and post-intervention (i.e., week 4) values of each group by paired t-test or Wilcoxon signed-rank test (*: p < 0.05; **: p < 0.01; the dashed line represents the median baseline value for the respective MSI), and (ii) compared the 4-week changes of the three interventions by one-way ANOVA or Kruskal-Wallis test (if the result was significant, a post-hoc Tukey-Kramer multiple comparison was performed to determine the significance of each group combination). Acupuncture was the only intervention that resulted in significant changes in the four MSIs (*: p < 0.05). The changes in MI and FI induced by Acu were significantly different from those by ShamAcu and NoAcu (MI: p = 0.040, one-way ANOVA; FI: p = 0.043, one-way ANOVA). Multiple comparisons showed that Acu resulted in a greater increase in MI than ShamAcu (^+^: p = 0.034) and a greater reduction in FI than NoAcu (^#^: p = 0.059).

We next examined longitudinal changes of the clinical scores and MSIs from week 0 to 4 by considering each group separately. Intriguingly, while for the 5 clinical scores every intervention group (Acu, ShamAcu, and NoAcu) showed significant improvement at week 4 (20 of 20 instances), for the MSIs improvements were significant only in a small subset of them (10 of 48 instances) (Table 2). Specifically, Acu induced significant changes in 5 MSIs (DevD_o_, MI, FI, BFRR_C_, DO), whereas ShamAcu and NoAcu, in only 0 and 2 MSIs (ITV_BFRR_C_, ITV_BFRR_C_ (mod)), respectively (Table 2).

When we examined the correlations between the 4-week change in MSIs and the 4-week change in clinical scores, we found that 19 of 192 correlations were significant (Fig. 4). Interestingly, for Acu, 9 of 48 instances were statistically significant, with the pre-to-post direction of MSI changes for all 9 instances being towards the normative as the clinical scores improved. For ShamAcu and NoAcu, only 3 and 2 instances were significant, respectively (Fig. 4). Specifically, for the 3 significant correlation instances in ShamAcu, DevD_O_, DevD_A_, and FI changes were positively correlated with changes in clinical scores instead of negatively correlated. Since the values of stroke survivors were greater than the baseline values for these 3 MSIs, the increase in clinical scores induced by ShamAcu was associated with these MSIs further deviating from their normative levels. This observation suggests that compensation probably constitutes a considerable portion of the motor recovery induced by ShamAcu, consistent with the above finding that improvement in clinical scores in ShamAcu was not accompanied by the corresponding recovery of MSIs.

**Figure 4.**
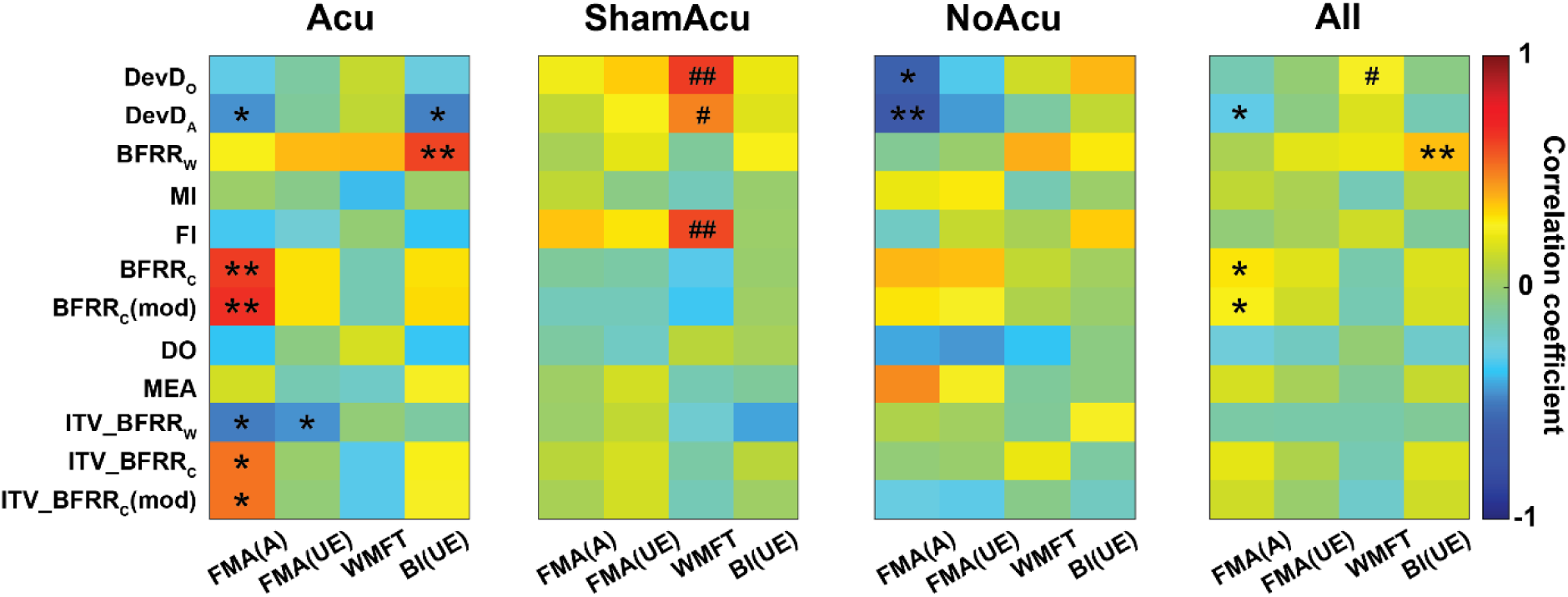
Pearson correlation between the 4-week change in muscle synergy indexes (MSIs) and the 4-week change in clinical scores for the Acu group (n = 21), ShamAcu group (n = 21), NoAcu group (n = 17), and all RCT participants (n = 59). While in many instances, the increase in clinical scores was associated with MSIs changing towards the normative (*: p < 0.05; **: p < 0.01), in some instances, the increase in clinical scores was associated with MSIs further deviating from the normative (^#^: p < 0.05; ^##^: p < 0.01).

Overall, the effects of Acu could be better differentiated from those of ShamAcu and NoAcu with MSI changes or correlations between MSI and clinical score changes than with clinical score changes alone. Acu appears to be better able to improve clinical scores through restorations of more MSIs than ShamAcu and NoAcu.

### Subjects assigned by MSI predictive models had greater recovery

To investigate whether the pre-intervention MSIs have the potential to predict rehabilitation outcomes, we computed the Pearson correlation between each MSI at week 0 and the absolute change in each clinical score (Fig. 5A) or its realized recovery (Fig. 5B) at week 4, with each MSI considered individually. We found that MSIs were better at predicting the realized recovery, as a greater proportion of correlations were significant for the realized recovery (48 of 192 instances) than for the absolute score change (24 of 192).

**Figure 5.**
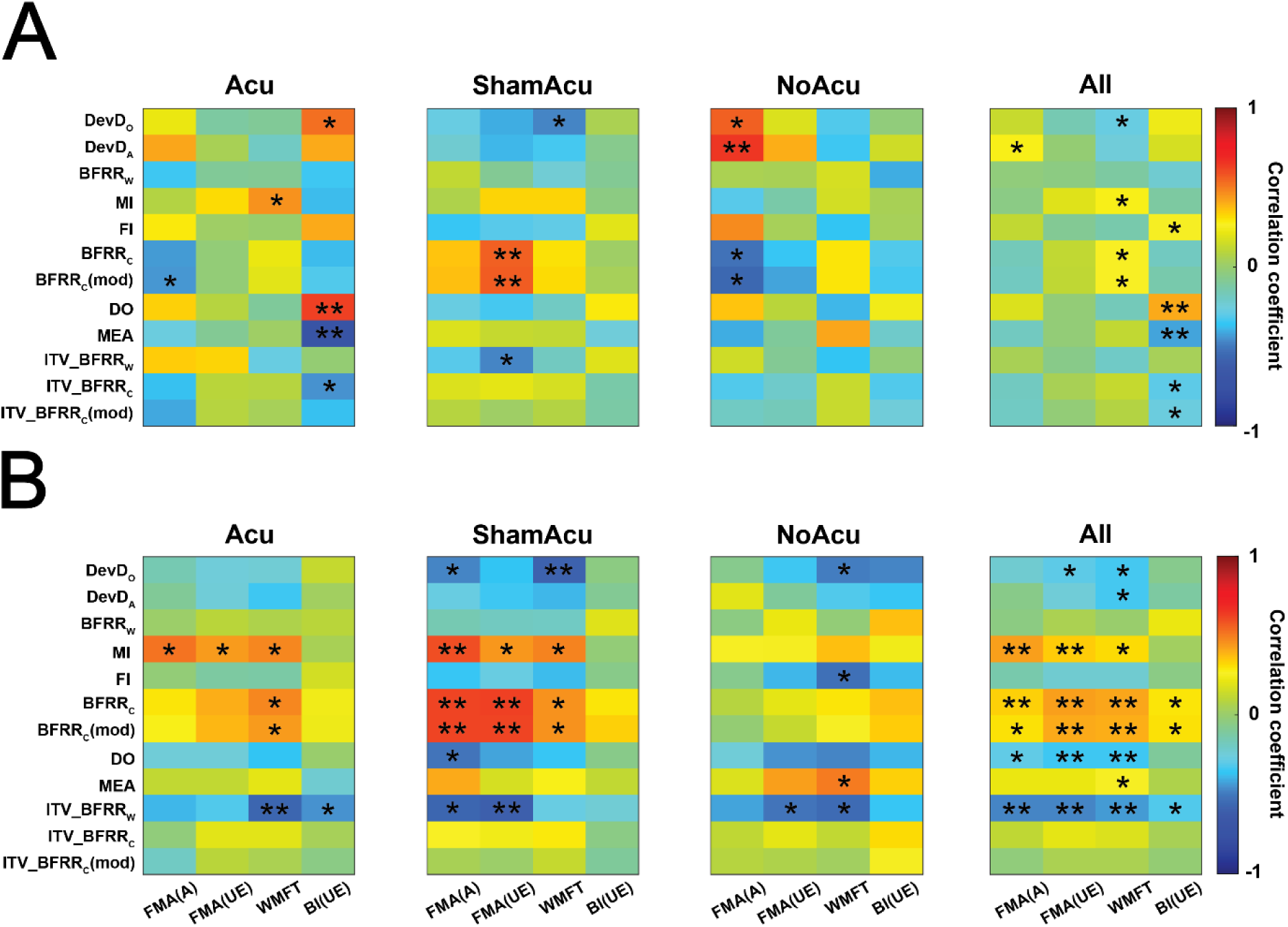
Predictive values of individual muscle synergy indexes. (A) Pearson correlation between muscle synergy indexes (MSIs) at week 0 and the 4-week change in clinical scores for the Acu group (n = 21), ShamAcu group (n = 21), NoAcu group (n = 17), and all RCT participants (n = 59). (B) Pearson correlation between MSIs at week 0 and the 4-week realized recovery of clinical scores for the same four groups. *: p < 0.05; **: p < 0.01.

The above results prompted us to perform a stepwise multiple linear regression to systematically select variables from the MSIs, clinical scores, and other clinical data (i.e., age, gender, affected side, and post-stroke duration) as pre-treatment inputs to predict the realized recovery of clinical scores (Supplementary Table 7). To illustrate, Fig. 6A shows the predictive models for FMA(A). The in-sample R^2^ was relatively low when we constructed overall predictive models for all intervention groups (median R^2^ = 0.34), but became higher when we constructed separate predictive models for each intervention group (median R^2^ = 0.51), the smaller sample sizes notwithstanding. Moreover, the combination of MSIs and clinical scores as inputs yielded R^2^ values (0.41 ± 0.21) that were considerably higher than clinical scores alone (0.26 ± 0.21).

**Figure 6.**
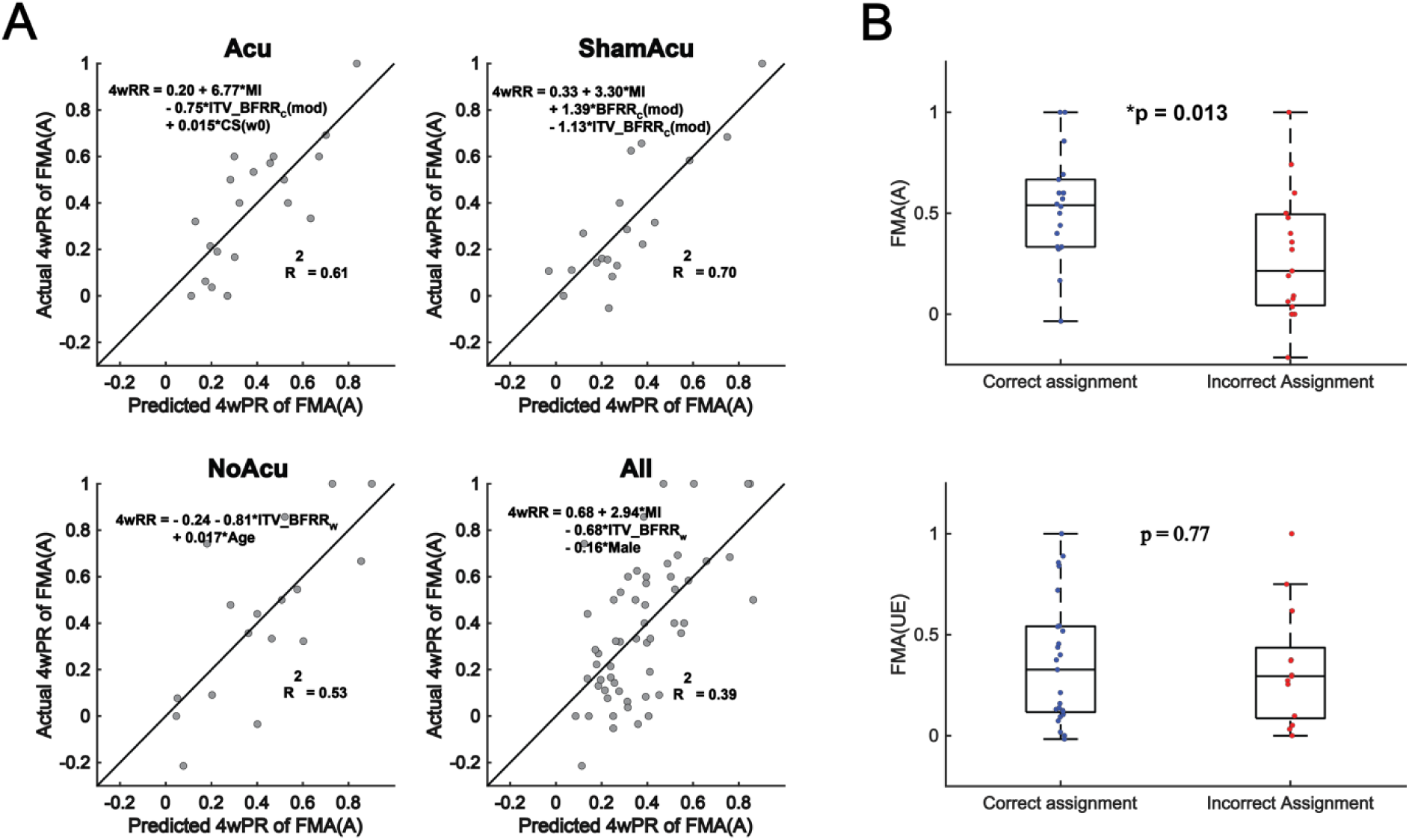
Predictive models derived from muscle synergy indexes (MSIs) and clinical data. (A) Predictive models for 4-week realized recovery of FMA(A) developed for the Acu group (n = 20), ShamAcu group (n = 19), NoAcu group (n = 17), and all RCT participants (n = 56), using stepwise multiple linear regression. (B) A simulation of the classification process in retrospect. We calculated the expected realized recovery of FMA(A) or FMA(UE) of each stroke survivor who received acupuncture or no acupuncture to determine whether he/she was correctly assigned to the right intervention. After that, the actual realized recovery of FMA(A) or FMA(UE) was compared between the correctly assigned and the incorrectly assigned (FMA(A): *p = 0.013, independent t-test; FMA(UE): p = 0.77, Mann-Whitney U test).

We evaluated the applicability of the predictive models in determining which stroke survivors were the most appropriate candidates for each intervention group by performing a simulation of the classification process in retrospect. To illustrate, we consider the stroke survivors who received Acu (n = 21) and NoAcu (n = 17), respectively. First, for every member of both groups, we calculated both the expected realized recovery after acupuncture and no acupuncture, respectively. For each Acu member, we calculated the expected realized recovery after acupuncture by using the predictive model based on the data of all other Acu members (i.e., analogous to leave-one-out cross-validation), and that after no acupuncture, by using the predictive model developed from the NoAcu group (i.e., analogous to external validation). We calculated the expected realized recovery after acupuncture and no acupuncture for every NoAcu subject analogously. Second, we determined the optimal intervention group for each Acu and NoAcu member by choosing the intervention with the larger expected realized recovery. Third, we classified each Acu and NoAcu member as being correctly or incorrectly assigned to his/her intervention group, as follows. A stroke survivor was deemed correctly assigned if he/she actually received the intervention determined by the predictive models to yield the larger expected recovery, and incorrectly assigned otherwise. Finally, we determined the validity of the predictive models and our classification by comparing the actual realized recovery of the correctly and incorrectly assigned subjects.

The above procedure was performed for FMA(A) and FMA(UE). For both scores, the mean or median actual realized recovery was higher in the correctly assigned group than in the incorrectly assigned group, but the difference was significant only for FMA(A) (t(35) = 2.50, p = 0.013) but not for FMA(UE) (z = 0.29, p = 0.77) (Fig. 6B).

## DISCUSSION

Muscle synergies may provide clinically valuable information for neurological disorders such as stroke because they are a relatively simple and compact representation of the coordinative structures utilized by the motor system to construct movement.^38^ Thus, muscle synergies may enable us to readily characterize and compare the motor functional statuses of diverse stroke survivors.^11^ Here, we designed 12 MSIs to quantify a wide variety of post-stroke characteristics of muscle synergies and their activation profiles (Fig. 1; Supplementary Note 4) and demonstrated how these MSIs could be used for motor functional assessment, clarification of the therapeutic effects of different interventions, and prediction of post-intervention motor outcome.

In the past decade, muscle synergy analysis has been progressively refined to investigate the pathophysiological basis of neurological disorders^16^ (Supplemental Table 1). However, previous studies were limited in that they focused more on the characteristics of spatial synergies (i.e., our MSI categories 1–3) (Supplementary Table 1) than temporal coefficients (i.e., MSI categories 4–5)^16^, and that only a subset of MSIs were shown to correlate with clinical scores due to the limited number of MSIs included or a relatively small cohort size. To overcome these limitations, we conducted a more comprehensive and robust muscle synergy analysis on our stroke survivors by including MSIs from all 6 categories and recruiting a much larger number of patients. The fact that all 12 MSIs in our study were significantly correlated with multiple clinical scores (Fig. 2) demonstrated the MSIs’ ability to assess the motor functional status of stroke survivors.

If muscle synergies directly reflect the organizational structures of the motor system,^38^ as has been suggested by recent studies on their neural basis,^9,39^ the use of MSIs for motor assessment may permit the distinction between true recovery and the emergence of compensation during motor recovery. Specifically, true recovery can be viewed as an increase in functional scores accompanied by the return of MSIs to their prestroke levels while compensation is functional improvement without the MSIs’ return. In our study, while all clinical scores in the Acu, ShamAcu, and NoAcu groups improved significantly, only a small subset of MSIs changed towards the normative after intervention (Table 2). Thus, acquiring compensatory strategies presumably plays a significant role in the recovery of our stroke survivors. This result is reminiscent of previous trials in which there was improvement only in the clinical scores but not kinematics,^40^ and consistent with the fact that basic rehabilitation therapies generally focus more on successful task completion but less on movement quality or the movement’s underlying muscle patterns.^41^

While compensation could result in immediate functional gain, it is less desirable than true recovery in terms of long-term outcomes.^22,40,41^ Recently, a number of new personalized therapies have been proposed to specifically target the true recovery of muscle synergies, some of which may be used in conjunction with conventional therapies.^11,13,42^ Since the 12 MSIs in our study can effectively characterize impairment-relevant muscle synergy changes after stroke (Fig. 2), the synergy features they describe may themselves be potential therapeutic targets for future synergy-based therapies.

As an adjunctive rehabilitative therapy, abdominal acupuncture appears capable of facilitating alterations and restorations of muscle synergies (Fig. 3). When we performed Pearson correlation on changes in MSIs with changes in clinical scores, Acu, but not ShamAcu nor NoAcu, showed 9 correlations with improvement of the clinical scores accompanied by changes of 7 MSIs towards their normative levels (Fig. 4). Also, when we considered just MSI changes of individual groups, while ShamAcu and NoAcu induced significant changes in 0 and 2 MSIs, respectively, Acu induced significant changes in 5 MSIs (DevD_O_, MI, FI, BFRR_C_, and DO; Table 2). Of the 5, 4 changed towards the normative, while MI deviated further away from the baseline (Fig. 3). In our multi-group comparisons, Acu was more effective than ShamAcu and NoAcu in promoting the recovery of FI and MEA, but it also induced more merging than the other interventions. Overall, these results indicate that acupuncture may facilitate true recovery of some MSIs, but it may also permit stroke survivors to adopt compensatory strategies along specific dimensions, such as merging. This analysis highlights how the MSIs may clarify the effects of different rehabilitative options in a clinical trial. Notably, the Acu, ShamAcu, and NoAcu groups studied here would have shown minimal between-group differences had only the clinical scores been adopted as the outcome measures.

Despite the fact that among the three groups only Acu promoted more notable changes in some MSIs, we did not observe any significant intergroup differences in the post-intervention changes in clinical scores, except that Acu showed more improvement of score BS at week 2. In fact, all groups showed some positive changes in all scores by week 4 (Table 2). One possibility is that the positive score changes in the three groups are achieved via different patterns of MSI changes. As noted above, score recovery in Acu may overall be due more to the true recovery of the MSIs (though compensation was not excluded), while those in ShamAcu and NoAcu, more to the acquisition of compensation. This interpretation is reasonable, especially given that in ShamAcu, positive changes in clinical scores were associated with some MSIs (i.e., DevD_O_, DevD_A_, and FI) becoming further deviated from their normative levels (Fig. 4). Another possibility is that the therapeutic potential of abdominal acupuncture, as reflected by the clinical scores, is underestimated in this trial because it yields maximal benefit only in a subgroup of stroke survivors. After randomized subject assignment, the less favorable clinical outcomes of the incorrectly assigned subjects may have obscured the favorable outcomes of the correctly assigned. It is plausible that acupuncture modifies muscle synergies in a manner that is only advantageous for those with particular pre-intervention characteristics. For example, Naeser et al.^30^ reported that acupuncture was only effective if the lesion in the motor areas and pathways was less than 50%.

Since many pre-treatment MSIs individually correlated with the realized recovery of clinical scores (Fig. 5), we tested the feasibility of using multiple MSIs as a predictive instrument to determine which stroke survivors would or would not benefit most from acupuncture. In particular, we used MSIs, clinical data and stepwise multiple linear regression to develop intervention-specific predictive models for Acu and NoAcu, respectively, and used them to calculate the expected realized recoveries of both treatment options for each stroke survivor. As a proof of concept, our retrospective analysis based on assigning each subject to the treatment option with the best expected post-treatment outcome demonstrates that our predictive models are able to identify the option that could lead to a greater realized recovery in FMA(A) (Fig. 6).

Previous studies have relied on neuroimaging biomarkers to stratify stroke survivors into categories with different expected post-rehabilitation potentials.^4,5^ Here, we demonstrate that MSIs extracted from EMG-derived muscle synergies may likewise predict post-treatment functional outcomes (Fig. 5; Fig. 6). The MSIs may therefore be used as alternative recovery markers that are potentially easier and less expensive to obtain once the process of measuring the MSIs is further streamlined.^43^ Our study is also unique in that we explore the outcome prediction of multiple treatment options – usual care with or without acupuncture – through different intervention-specific predictive models. Prior studies have focused on predicting the recovery potential of a single rehabilitation option, such as usual care^23,35,44–47^ and robotic therapy.^48^ With this prediction, clinicians may then suitably customize the goals and parameters of that particular treatment based on the anticipated potential for the best final outcome. Here, by making predictions on multiple treatment options, it becomes possible to identify the optimal option for every stroke survivor by comparing the expected outcomes of all available interventions. As more intervention-specific predictive models are developed in the future, the overall efficacy of rehabilitative intervention will also increase when treatments are more precisely prescribed based on data-driven predictive models.

Our study is not without limitations. First, since we recorded EMGs primarily from shoulder and upper arm muscles, it is not unexpected that the MSIs were more closely related to FMA(A) than FMA(UE), with the former reflecting shoulder and elbow functions. Indeed, the absolute values of the correlation coefficients of MSIs with FMA(A) at week 0 were significantly greater than those with FMA(UE) (paired t-test, t(11) = 2.88, p = 0.015). Second, although many previous studies found a decreased number of synergies in severely impaired stroke survivors,^14,18,49–51^ we found an increase in dimensionality instead. This may be because our cohort was mostly in the early subacute phase of recovery, while the previously cited studies mostly recruited chronic patients. In fact, a recent study that analyzed both subacute and chronic stroke survivors also found an increased mean dimensionality.^52^ Moreover, merging is likely a compensatory strategy for increased dimensionality, adopted by stroke survivors at an early stage of recovery. While healthy individuals exhibit a minimal degree of merging by definition, a significantly higher level of merging was observed among our stroke survivors (p < 0.01), especially those with milder impairments (Figure 2). Considering that the number of muscle synergies increased in many of our stroke survivors after stroke, merging could have lessened their computational burden by reducing the already increased dimensionality of motor control. Since muscle synergies may evolve at different stages through merging and fractionation,^14^ future studies could consider tracking the muscle synergies of stroke survivors from acute to chronic phases of recovery. Third, since this is a single-center study, the lack of diverse ethnic groups may be a limitation.

## CONCLUSION

Muscle synergy indexes (MSIs) are recovery biomarkers that reflect the control architecture of the motor system. In this study, we have formulated 12 MSIs and demonstrated in a clinical trial of acupuncture how these MSIs could be used to evaluate the motor functional statuses of stroke survivors, clarify the effects of different interventions, and predict the motor outcomes of multiple rehabilitative options. Our results directly support that EMG-derived muscle synergy could be a useful biomarker in neurorehabilitation. How muscle synergies may be best employed in conjunction with other neurological biomarkers to derive more accurate outcome predictions is a direction that awaits future research.

## Acknowledgements

We thank The CUHK School of Biomedical Sciences and The Guangdong Provincial Hospital of Traditional Chinese Medicine for their support of this study, and Prof. Lu Gang for his administrative assistance and support.

## Data Availability Statement

All data reported in this study as well as the codes developed for the analyses are available to any researcher upon any reasonable request sent to the corresponding authors.

